# Targeted ERAS implementation for postoperative care after Bellwether procedures in Africa: A pragmatic cluster-randomized trial from Ethiopia

**DOI:** 10.1101/2025.11.18.25339361

**Authors:** Fitsum Kifle Belachew, Peniel Kenna Dula, Ermiyas Belay Woldesenbet, Betelehem Mulye, Desta Galcha, Kalkidan Kifle, Dagmawi Dagne, Megbar Dessalegn Mekonnen, Kokeb Desta Belihu, Tewodros Kifleyohannes, Brook Demissie, Abiy Dawit, Salome Maswime, Bruce Biccard, NaPQIN ERAS trial collaborators

## Abstract

**Introduction:** In Africa, where access to timely and safe surgical care remains limited, postoperative complications and prolonged hospital stays continue to challenge health systems. The Enhanced Recovery After Surgery (ERAS) protocol has been shown to improve perioperative outcomes by reducing hospital length of stay (LOS) and complications, but compliance remains inconsistent.

**Objective:** To determine whether improving ERAS compliance in Ethiopia, through a “Triple Intervention Strategy” of early postoperative feeding, ambulation, and urinary catheter removal, could reduce hospital LOS for patients undergoing laparotomy and cesarean section (CS).

**Methods:** This study was designed as a cluster-randomized clinical trial conducted across 10 hospitals within the National Perioperative Quality Improvement Network (NaPQIN) in Ethiopia. Hospitals were randomly assigned to either the intervention group (n=5), which received structured ERAS training reinforced through continuous monitoring and supervision, or the control group (n=5), which continued standard perioperative care without additional reinforcement. The primary outcome was hospital LOS, and secondary outcomes included compliance with the ERAS components, determinants of LOS, and postoperative complications. Data were managed through the NaPQIN platform and analyzed using R statistical software.

**Results:** A total of 8,256 patients were enrolled, with 5,887 (71.3%) in the intervention group and 2,369 (28.7%) in the control group. Full compliance with the ERAS bundle improved to 76.5% in the intervention group compared to 57.9% in controls (p < 0.001). Patients in the intervention group had a significantly shorter LOS (mean 80.75 vs. 89.24 hours; p < 0.001). The intervention group also had significantly fewer postoperative complications (2.1% vs 4.8%; p < 0.001), and more patients were discharged without any complications.

**Conclusions and Relevance:** This pragmatic trial, enabled by a national perioperative data system, demonstrated that the targeted implementation of postoperative ERAS elements, early oral feeding, mobilization, and timely urinary catheter removal significantly improved compliance and reduced hospital stay without requiring additional resources. While full ERAS pathways remain the ideal, focused, context-adapted strategies can offer scalable benefits in LMIC settings burdened by surgical backlogs and limited perioperative capacity. Broader adoption should prioritize tailored integration, ongoing evaluation, and provider engagement to maximize system-wide impact.

*Trial Registration:* pactr.samrc.ac.za identifier **<u>PACTR202502863551536</u>**

## Background

Surgical care is a vital component of public health, as evidenced by the Lancet Commission on Global Surgery, Disease Control Priorities 3, and World Health Assembly resolution 68.15 (1). Between 2004 and 2012, major surgical procedures increased from 234.2 million to 312.9 million per annum (2). Despite the increase in surgical volume, adverse outcomes, including postoperative complications and mortality, remain significant concerns globally (3). Africa, home to many low-and middle-income countries (LMICs), has unique challenges in surgical healthcare, with opportunities to improve postoperative mortality and complications, currently reported at 2.1%, and 18.2%, respectively, despite lower surgical volumes and healthier patients than global comparators (4). Ethiopia exemplifies the challenge of delivering surgical care in a low-resource environment (5). The number of surgeries performed in Ethiopia’s public health facilities falls significantly short of the target of 5,000 surgeries per 100,000 population annually. This situation highlights the need for quality care initiatives such as the Enhanced Recovery After Surgery (ERAS) to enhance surgical care and improve patient outcomes (6).

ERAS is a multimodal approach to perioperative care that combines evidence-based practices, including preoperative counselling, minimally invasive surgical techniques, and early mobilisation, to optimise patient outcomes, reduce complications, and expedite recovery, thereby shortening hospital stays following surgery (7). Although research is limited, a systematic review and meta-analysis of gastrointestinal surgery suggests that ERAS practices in African settings offer similar benefits to those in high-income countries, although the dataset is small (8). These benefits include shorter hospital stays, reduced complications, and improved patient outcomes, which are crucial in resource-constrained settings to minimize surgery-related costs (8,9). A study of a perioperative registry in Ethiopia assessed compliance with ERAS protocols and found a 65.2% compliance rate for Bellwether surgeries (10). This suggests a significant opportunity to improve ERAS implementation in some LMICs to improve surgical outcomes.

In this trial, we aimed to determine whether improving ERAS compliance in Ethiopia through a bundled “Triple Intervention Strategy” of early postoperative feeding, ambulation, and urinary catheter removal for patients undergoing laparotomy and cesarean section would decrease hospital stay.

## Material and Methods

This trial adheres to the 2010 Consolidated Standards of Reporting Trials (CONSORT) extension for cluster-randomized studies (11). The trial protocol is registered in the Pan-African Clinical Trials Registry (*PACTR202502863551536*) (12). Ethical approval was obtained from the University of Cape Town Faculty of Health Sciences Human Research Ethics Committee (*Protocol Number: 220/2024*) and the Arba Minch University Institutional Research Ethics Review Board Office (*Protocol Number: IRB/2320/2024*). Given that the study recommends strict compliance with the available guidelines and evaluates clinical practice patterns, the ethics committees waived the requirement for written informed consent. All data were de-identified and securely stored to ensure confidentiality.

### Trial design

This study was a pragmatic, cluster-randomized clinical trial designed to evaluate the effectiveness of a structured training intervention on ERAS protocol compliance. The study was conducted across 10 hospitals participating in the National Perioperative Quality Improvement Network (NaPQIN), a collaborative platform supporting hospitals in implementing evidence-based perioperative practices, monitoring outcomes, and facilitating quality improvement initiatives (13).

Hospitals were randomly assigned to either the intervention group (n = 5) or the control group (n = 5) using a computer-generated allocation sequence in R, conducted by an independent statistician. No stratification was applied in the randomization process.

The intervention group received structured ERAS training on implementing the intervention, as defined by the LMICs’ ERAS guidelines (14). Continuous reinforcement strategies included routine monitoring via the NaPQIN platform, supervision by designated site investigators, broadcast messages in recovery rooms and wards, and regular feedback and evaluation meetings. The control group received standard perioperative care without training or reinforcement interventions.

### Participants

Eligibility included patients 18+ undergoing laparotomy or cesarean sections (CS) who consented. Exclusions were intraoperative deaths, ICU admissions or referrals post-surgery, procedures other than laparotomy or CS, or patients under 18. Recruitment from January to December 2024 involved tracking all registered patients’ surgical interventions and outcomes on the NaPQIN platform.

### Intervention

Hospitals were randomized at the cluster level into either the intervention or control group using a computer-generated allocation sequence prior to the training. Following randomization, all participating hospitals in the intervention group received ERAS training as part of the NaPQIN initiative, ensuring a standardized approach to perioperative care.

### Training program

The ERAS training sessions, conducted by eight trainers who completed a ToT program, took place from December 6-9, 2023. The training included general surgeons, gynecologists, obstetricians, anesthesia professionals, and nurses from PACU, surgical wards, and gynecology departments. It emphasized safe OR practices and ERAS, with the Safe OR course developed to provide multidisciplinary training for safe surgery, uniting various perioperative healthcare providers who rarely train together (15). Launched in 2017, the course began as a three-day program aimed at improving surgical safety and team-based perioperative management. Although it briefly introduced ERAS principles, it did not fully cover their clinical implementation. For this study, the course was expanded to four days to include detailed ERAS protocols, NaPQIN platform, guidelines, recommendations, and strategies. It also retained core modules from the original Safe OR course on teamwork, safety, decision-making, patient optimization, anesthesia, analgesia, emergency management, recovery, and quality improvement. These sessions built a solid knowledge base for participating in hospitals.

### Reinforced implementation in the intervention group

Following training, the five hospitals assigned to the intervention group implemented a reinforced ERAS compliance strategy. To ensure structured compliance, lead hospital investigators were designated at these sites to oversee implementation and reinforce adherence.

The surgical team played a central role in leading protocol implementation, ensuring alignment between different perioperative teams, and promoting compliance with ERAS principles as part of routine patient care.

### Control group

The five hospitals assigned to the control group followed standard perioperative care practices without additional reinforcement strategies beyond the initial training.

### Outcomes

The primary outcome was hospital LOS, measured in hours from surgical admission until hospital discharge. Secondary outcomes included: i) compliance to ERAS protocol components, including early feeding, urinary catheter removal, and early ambulation, ii) factors influencing LOS, including patient demographics (age, ASA classification, urgency of surgery), surgical characteristics (procedure type, duration), and cluster-level variability, and iii) postoperative complications assessed using the Clavien-Dindo complication score system.

Data were collected from the NaPQIN platform, allowing for standardized, real-time data reporting across all study sites.

### Sample size

A *priori* sample size was determined using the Donner & Klar sample size formula to estimate the effect size for a continuous outcome (16). We proposed an ICC (intra-cluster correlation) of 0.02 based on the general recommendations due to the uncertainty of the correlation coefficient in epidemiologic studies (17). We determined that 1,000 patients (500 per arm) were required to detect a clinically meaningful difference in hospital length of stay, defined as a moderate standardized effect size of 0.4 (48-hour reduction in length of stay), with an alpha value of 5% and 80% power. Considering the estimated number of samples per cluster is 100, and assuming equal sizes across clusters, we enrolled 5 clusters per arm, totaling 500 patients per group and 1000 in total. We used the sample size determination formula coded in R(18). This function uses a “while loop” in R to adjust the number of clusters per arm iteratively, K_1_(set as zero as a starting point), until a stable value k is reached. The t-distribution was used instead of the Z-distribution. Although the minimum required sample size was 1,000, we included all eligible participants identified during the study period, resulting in a final sample of 8,256 patients.

### Blinding

Blinding was not feasible for investigators and healthcare providers due to the broadcast messages, reinforcement strategies, and direct involvement in ERAS compliance monitoring. However, statistical analyses were conducted with blinding, ensuring that the statistician remained unaware of group allocation to prevent bias in data interpretation.

### Statistical analysis

Descriptive and inferential statistical approaches were undertaken to assess factors influencing LOS and evaluate the differences between the control and intervention groups. Descriptive statistics were used to summarize baseline characteristics, with frequencies and percentages for categorical variables, while means (medians) with standard deviations (interquartile ranges) were used for continuous variables as appropriate. Group-level comparisons were conducted using the Chi-square test of independence, t-test, or ANOVA, as applicable, based on the type of variables compared.

The extent of missing data and its distribution were evaluated. Accordingly, we applied multiple imputations that considered the clustering effect on missingness by study clusters using Multiple Imputation with Chained Equations (MICE). After performing the imputation, the data generally preserves the pre-imputation data distribution in both categorical and numerical variables. To reduce the influence of extreme outliers, winsorization was made at the 10th and 90th percentiles prior to imputation. The normality assumption was checked using the Kolmogorov–Smirnov test. For any deviation from the normality assumption, a non-parametric test was applied to compare the differences between groups.

To estimate the effect of intervention while adjusting for potential confounders, Generalized Estimating Equations (GEE) with an exchangeable correlation structure were used. This approach accounted for the clustering of patients within hospitals, adjusting for age, surgical type, educational level, urgency of surgery, and intravenous (IV) fluid volume. We have used a residual versus fitted values plot to assess model adequacy. Accordingly, a slight curvature was noted, indicating potential mild nonlinearity; however, the overall pattern suggests an acceptable model fit. Moreover, we have provided the log-transformed version of the model as a supplementary file.

In addition to individual patient-level analysis, cluster-level analyses were conducted to explore variability within and between hospitals. Ranked cluster means were used to provide further insight into group-level differences, highlighting institutional variations in ERAS adherence and LOS outcomes. All statistical analyses were performed using R statistical software version 4.4.2.

## Results

### Baseline characteristics

A total of 8,256 patients were recruited for the study from January to December 2024 (as shown ***in* *Figure 1***), comprising 5,436 (65.8%) who underwent cesarean section (CS) and 2,810 (34.2%) who underwent laparotomy. The majority of patients were males, at 1,482 (52.7%). Of these, 5,887 (71.3%) were in the intervention group, while 2,369 (28.7%) were in the control group. There were differences between the two groups across all baseline characteristics (p < 0.001). Most procedures were non-scheduled, accounting for 6,759 (81.9%) of all surgeries, including 77.7% in the control group and 71% in the intervention group. All patients were classified as ASA I or II. Differences were observed across age, marital status, education, occupation, surgery urgency, and ASA class, as shown in **Table 1**.

**Figure 1:**
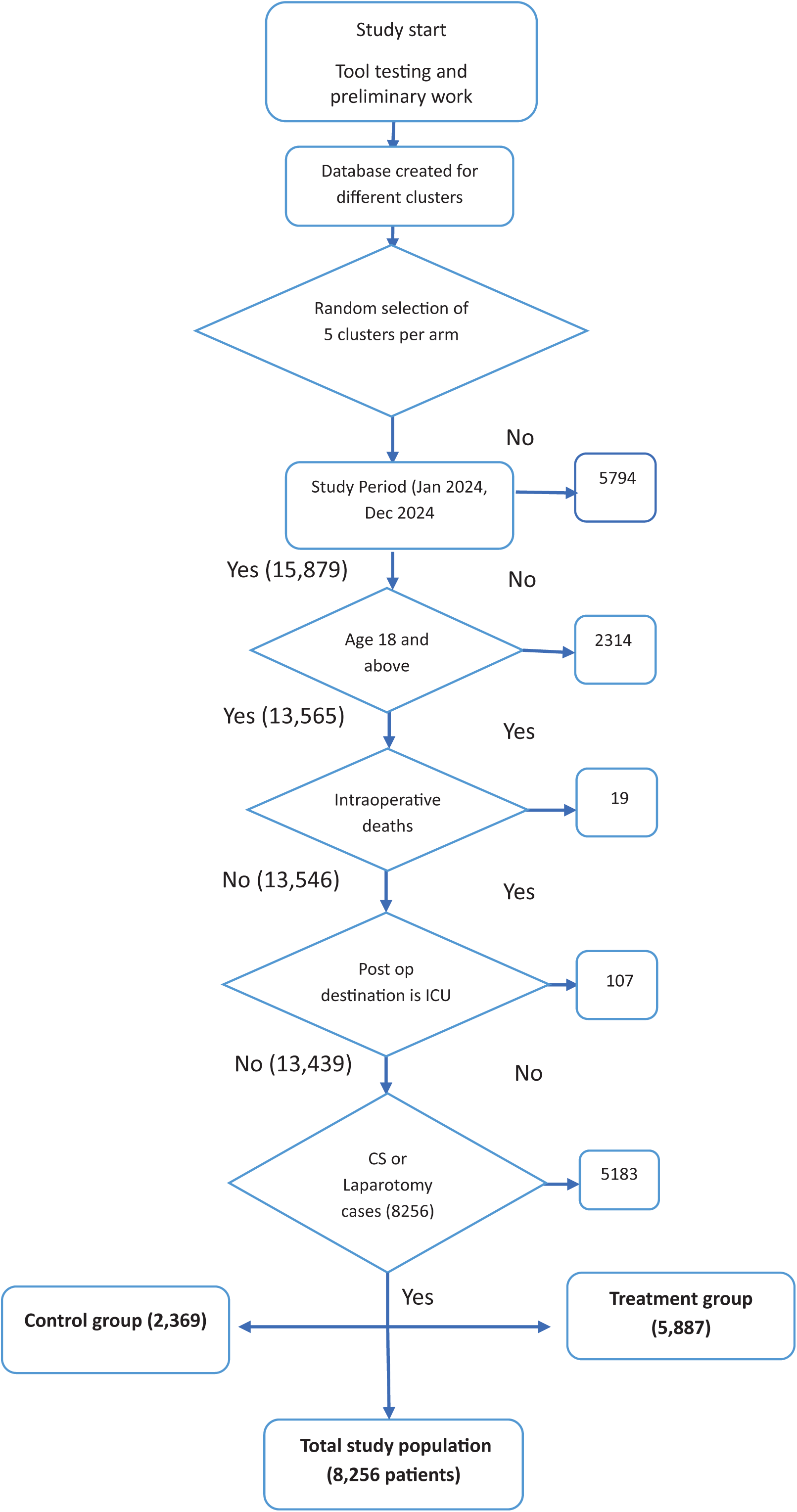
CONSORT diagram: screening, exclusion, and final study population.

**Table 1:** Baseline characteristics of study participants; the intervention and control groups.

| Characteristic | Control group, N = 2369 |  |  |  | Intervention group, N = 5887 |  |  |  |
| --- | --- | --- | --- | --- | --- | --- | --- | --- |
|  | N | CS<br>N = 1,559<sup>1</sup> | Laparotomy<br>N = 810<sup>1</sup> | p-value<sup>2</sup> | N | CS<br>N = 3,877<sup>1</sup> | Laparotomy<br>N = 2,010<sup>1</sup> | p-value<sup>2</sup> |
| Age group | 2,369 |  |  | <0.001 | 5,887 |  |  | <0.001 |
| 21-30 |  | 1,152 (74%) | 329 (41%) |  |  | 2,969 (77%) | 775 (39%) |  |
| 31-40 |  | 394 (25%) | 172 (21%) |  |  | 883 (23%) | 462 (23%) |  |
| 41-50 |  | 13 (0.8%) | 309 (38%) |  |  | 25 (0.6%) | 773 (38%) |  |
| Sex | 2,369 |  |  | <0.001 | 5,887 |  |  | <0.001 |
| Male |  | 0 (0%) | 434 (54%) |  |  | 0 (0%) | 1,048 (52%) |  |
| Female |  | 1,559 (100%) | 376 (46%) |  |  | 3,877 (100%) | 962 (48%) |  |
| Marital status | 2,369 |  |  | <0.001 | 5,887 |  |  | <0.001 |
| Married |  | 1,530 (98%) | 584 (72%) |  |  | 3,728 (96%) | 1,501 (75%) |  |
| Single |  | 17 (1.1%) | 208 (26%) |  |  | 32 (0.8%) | 392 (20%) |  |
| Other |  | 12 (0.8%) | 18 (2.2%) |  |  | 117 (3.0%) | 117 (5.8%) |  |
| Education level | 2,369 |  |  | <0.001 | 5,887 |  |  | <0.001 |
| Illiterate |  | 138 (8.9%) | 132 (16%) |  |  | 571 (15%) | 401 (20%) |  |
| Elementary |  | 254 (16%) | 209 (26%) |  |  | 903 (23%) | 400 (20%) |  |
| Highschool |  | 578 (37%) | 213 (26%) |  |  | 1,060 (27%) | 682 (34%) |  |
| College and above |  | 549 (35%) | 235 (29%) |  |  | 897 (23%) | 241 (12%) |  |
| Other |  | 40 (2.6%) | 21 (2.6%) |  |  | 446 (12%) | 286 (14%) |  |
| Occupation | 2,369 |  |  | <0.001 | 5,887 |  |  | <0.001 |
| Government employed |  | 205 (13%) | 95 (12%) |  |  | 410 (11%) | 93 (4.6%) |  |
| Private work |  | 621 (40%) | 178 (22%) |  |  | 710 (18%) | 576 (29%) |  |
| Student |  | 37 (2.4%) | 92 (11%) |  |  | 27 (0.7%) | 176 (8.8%) |  |
| Housewife |  | 547 (35%) | 173 (21%) |  |  | 1,494 (39%) | 314 (16%) |  |
| Other |  | 149 (9.6%) | 272 (34%) |  |  | 1,236 (32%) | 851 (42%) |  |
| Urgency of Surgery | 2,369 |  |  | <0.001 | 5,887 |  |  | <0.001 |
| Scheduled |  | 311 (20%) | 217 (27%) |  |  | 874 (23%) | 815 (41%) |  |
| Non-scheduled |  | 1,248 (80%) | 593 (73%) |  |  | 3,003 (77%) | 1,195 (59%) |  |
| ASA Class | 2,369 |  |  | <0.001 | 5,887 |  |  | <0.001 |
| I |  | 249 (16%) | 449 (55%) |  |  | 1,175 (30%) | 1,286 (64%) |  |
| II |  | 1,310 (84%) | 361 (45%) |  |  | 2,702 (70%) | 724 (36%) |  |

<sup>1</sup> = n (%)
<sup>2</sup> = Pearson's Chi-squared test
**ASA**- American Society of Anesthesiologist
**ERAS**- Enhanced Recovery After Surgery,
**NaPQIN**- National Perioperative Quality Improvement Network

Stratified comparisons by procedure showed no age or sex differences between intervention and control groups in CS and laparotomy cohorts (p > 0.05). Missing data across baseline variables was under 6% and was addressed with multiple imputations.

### Compliance with the ERAS interventions in practice

Full compliance with all three ERAS components was observed in 4,507 (76.5%) patients in the intervention group, compared to 997 (42.1%) in the control group (p < 0.001), as shown in ***Table 2***. For compliance with each component, whether alone or combined, 5,196 (87.0%) of patients in the intervention group complied with early mobilization compared to 1,482 (61.0%) in the control group (p < 0.001). Early feeding was achieved in 5,231(96.2%) of the intervention group compared with 1,519 (64.2%) of the control group (p < 0.001). Compliance with catheter removal was 5,489 (93.2%) of the intervention group and 1,883 (79.5%) of the control group (p < 0.001).

**Table 2:** Adherence rate to the recommended ERAS triple elements.

| Treatment |  | Intervention group<br>(N = 5887) | Control group<br>(N = 2369) | Overall<br>(N =8256) | P-value<br>(Chi-square) |
| --- | --- | --- | --- | --- | --- |
| **Early Mobilization** | Yes | 2774 (47.1%) | 1483 (62.6%) | 4257 (51.6%) | <0.001 |
|  | No | 3113 (52.9%) | 886 (37.4%) | 3999 (48.4%) |  |
| **Early Feeding** | Yes | 3242(55.1%) | 1509(63.7%) | 4751(57.6%) | <0.001 |
|  | No | 2645(44.9%) | 860(36.3%) | 3505(42.4%) |  |
| **Catheter Removal** | Yes | 4407(74.9%) | 1843(77.8%) | 6250(75.7%) | 0.009 |
|  | No | 1466(24.9%) | 526(22.2%) | 1992(24.1%) |  |
| **Combined triple ERAS elements** | Yes | 4507(76.5%) | 1372(57.9%) | 5879(57.9%) | <0.001 |
|  | No | 1380(23.5%) | 997(42.1%) | 2377(28.8%) |  |

ERAS- Enhanced Recovery After Surgery
N- Total population

### Hospital length of stay (LOS)

The overall hospital LOS was 83.19 (± 40.06) for patients in both groups. However, patients in the control group had a higher mean LOS of 89.24 hours compared to 80.75 hours in the intervention group (P<0.001), as shown in **Table 3**.

**Table 3:** Summary of the study participants’ hospital length of stay (LOS)

| Group | N | Mean | Median | SD | Min (OV) | Max (OV) | IQR | P-value |
| --- | --- | --- | --- | --- | --- | --- | --- | --- |
| **Overall** | 8,256 | 83.19 | 71.00 | 40.06 | 47.00 | 172.41 | 45.95 | **<0.001** |
| **Control group** | 2,369 | 89.24 | 72.08 | 44.02 | 47.00 | 172.41 | 63.27 |  |
| **Intervention group** | 5,887 | 80.75 | 70.00 | 38.08 | 47.00 | 172.41 | 44.13 |  |
| **Control vs intervention (T-test)** |  |  |  |  |  |  |  |  |

IQR- Interquartile range
Max - Maximum
Min -Minimum,
N - Number of participants,
OV- Observed values
SD - Standard Deviation,

### Postoperative complications and patient outcomes

The intervention group had fewer postoperative complications than the control group (2.1% vs 4.8%; p < 0.001), mostly of which were minor (Clavien-Dindo grade 1). Although patients in the intervention arm had a higher number of grade II and III complications, the overall rate of postoperative complications was significantly lowered in the intervention group as shown in **Table 4**.

**Table 4:** Postoperative complications by Clavien-Dindo classification across study groups.

|  |  | **Control group, N =**<br>**2369** | **Intervention group,**<br>**N = 5887** | **p-**<br>**value** |
| --- | --- | --- | --- | --- |
| No complication |  | 2,255 (95.2%) | 5,763 (97.9%) | <0.001 |
| With<br>Complication | 1 | 87 (3.7%) | 47 (0.8%) |  |
|  | 2 | 19 (0.8%) | 60 (1.0%) |  |
|  | 3 | 8 (0.3%) | 17 (0.3%) |  |

### Summary of factors influencing LOS

**Table 5** summarizes factors associated with hospital length of stay. Being in the intervention group was significantly associated with a shorter stay (RR = 0.921, 95% CI: 0.887–0.949; p < 0.001). Longer surgery duration, undergoing laparotomy (RR = 1.30), older age, and higher volumes of IV fluids were all associated with an increased length of stay. In contrast, higher education levels and elective surgery (RR = 0.944, p = 0.001) were related to a reduced length of stay.

**Table 5:** Factors influencing hospital length of stay description of intervention and control groups.

| Term | Coefficient | Std. error | RR | P value | 95% CI |
| --- | --- | --- | --- | --- | --- |
| **Intervention group** | -0.082 | 0.016 | 0.921 | <0.001 | (-0.113, -0.051) |
| Duration of surgery in minutes | 0.003 | 0.000 | 1.003 | <0.001 | (0.002, 0.004) |
| Surgery type - Laparotomy | 0.263 | 0.030 | 1.300 | <0.001 | (0.205, 0.321) |
| Education level - elementary | -0.038 | 0.015 | 0.963 | 0.011 | (-0.067, -0.009) |
| Education level - high School | -0.078 | 0.015 | 0.925 | <0.001 | (-0.107, -0.048) |
| Education level - college and above | -0.075 | 0.015 | 0.928 | <0.001 | (-0.105, -0.045) |
| Education level other | -0.121 | 0.026 | 0.886 | <0.001 | (-0.172, -0.071) |
| Age category (31-40 years) | 0.035 | 0.011 | 1.036 | 0.002 | (0.013, 0.057) |
| Age category (41-50 years) | 0.118 | 0.018 | 1.126 | <0.001 | (0.083, 0.154) |
| Total IV fluid (≤ 1000 ml) | 0.047 | 0.018 | 1.048 | 0.009 | (0.012, 0.082) |
| Total IV fluid (1000-1500 ml) | 0.059 | 0.021 | 1.061 | 0.006 | (0.017, 0.101) |
| Total IV fluid (1500-2000 ml) | 0.089 | 0.020 | 1.093 | <0.001 | (0.050, 0.128) |
| Urgency of surgery - non-elective | -0.057 | 0.018 | 0.944 | 0.001 | (-0.092, -0.023) |

CI - Confidence Interval, RR – Risk Ratio, Std. error - Standard error

## Discussion

This trial evaluated a bundled ERAS approach with early feeding, mobilization, and catheter removal across 10 Ethiopian hospitals. It reduced hospital stays, with intervention patients discharged 8.49 hours earlier (p < 0.001). Compliance with all three increased from 42.1% to 76.5% (p < 0.001). Full compliance among CS patients shortened LOS by 7.6 hours, and early catheter removal and feeding among laparotomy patients reduced LOS by 6.6 and 13 hours, respectively.

These results show that ERAS principles can be successfully adapted to low-resource settings without requiring additional infrastructure or personnel. The bundled ERAS strategy significantly increased compliance, aligning with previous findings that partial ERAS implementation shortens recovery time (10). However, compliance was lower among laparotomy patients, particularly regarding early mobility, likely due to factors such as increased pain, surgical invasiveness, longer surgical duration, exhaustion, and urgency of surgery, issues consistent with other studies (18–20). This may also have been impacted by inconsistent team coordination and a lack of standardized reinforcement strategies, which have been shown to affect ERAS compliance in other contexts (21,22). A broader systematic review also highlighted the heterogeneity of ERAS implementation in LMICs, often driven by gaps in supervision, training, and local adaptation (23). In addition to reducing LOS, the intervention group also had significantly fewer postoperative complications, with most being minor. This further supports the clinical benefit of targeted ERAS compliance, particularly in improving postoperative recovery.

This study implemented most of the ERAS Society’s postoperative care elements for LMICs, excluding multimodal analgesia due to resource limits (14). It successfully integrated early oral feeding, mobilization, and urinary catheter removal, with real-time audit via Ethiopia’s registry, aligning with ERAS’s focus on feasibility and system monitoring (14). Lessons from the African Surgical Outcomes Study (ASOS-2) trial showed the challenges of implementing complex surveillance in resource-limited settings (24). The study demonstrated that a simplified ERAS bundle can be effectively integrated into routine care, offering measurable benefits. Although ERAS guidelines target elective procedures, findings suggest selected elements may also improve outcomes in non-elective surgeries, broadening their applicability.

The digital feedback loop reinforced behavior change, improved fidelity, and fostered local accountability, while also enhancing outcome measurement accuracy. The low ICC of 0.029 indicated minimal between-cluster variation, supporting generalizability across hospitals. Though the reduction in LoS was small, scaled benefits include better bed turnover, fewer surgical backlogs, and lower inpatient costs in overburdened LMIC hospitals (25,26). These findings support that perioperative pathways like ERAS can cost-effectively strengthen health systems, especially where resources are limited (27). Multivariable analysis showed the intervention (RR = 0.921), elective surgery (RR = 0.944), and higher education linked to shorter LOS, while older age, laparotomy, longer surgeries, and more IV fluids predicted longer stays. These build on prior research by identifying modifiable practices and patient factors affecting recovery (10).

The 8.5-hour LOS reduction, smaller than the 52-hour average in a recent LMIC meta-analysis (23), was achieved through a simple, low-cost protocol. While earlier studies in LMICs, including a trial in Uganda and an Ethiopian audit, reported larger LOS reductions (18 to over 120 hours) using more intensive or broader perioperative strategies, our results show that a simplified ERAS bundle can still yield system-level benefits when contextually adapted into routine care (10,21). This supports evidence from high-income settings where higher protocol compliance correlates with shorter LOS and better outcomes (28). With support from stakeholders, digital tracking can help LMICs contribute to global ERAS initiatives (29).

While the trial aimed to include all eligible surgical patients across the 10 participating hospitals, several real-world factors affected patient recruitment and group balance. High surgical volumes, limited data entry capacity, documentation gaps, and intermittent service disruptions, such as anesthesia stockouts, autoclave malfunctions, backup generator failures, hospital renovations, and climate-related or seasonal events, contributed to missed cases, surgical cancellations, and inconsistencies in patient recruitment. These challenges may have introduced selection bias, impacted data completeness, and limited the generalizability of the findings across all LMIC hospital settings.

This pragmatic, registry-integrated trial, one of the few cluster-randomized ERAS studies conducted in LMICs, demonstrated that reinforcing key postoperative elements can significantly improve compliance and shorten hospital stays, even in the absence of additional infrastructure or staffing. While comprehensive ERAS adoption remains the ideal, prioritizing high-impact elements provides a feasible starting point for African health systems facing surgical backlogs and limited perioperative capacity. Future efforts should focus on multidisciplinary, hospital-wide ERAS integration, supported by continuous data feedback, routine training, and aligned policy frameworks. Increased investment and collaboration will be essential to scale these strategies and assess their long-term clinical and health system impact.

## Supporting information

Appendix 1_ERAS trial collaborators

## Data Availability

All data produced in the present study are available upon reasonable request to the authors

## Acknowledgments

This research was partially funded by the NIHR (NIHR133850), using the UK international development funding from the UK Government to support global health research and a seed grant funded by D-SINE Africa through its U54 project funds under Award Number U54TW012087, of the National Institutes of Health and of the Fogarty International Center and Office of Strategic Coordination (OSC). The views expressed in this publication are those of the authors and not necessarily those of the NIHR, the UK government, or any other funding bodies mentioned.

## Ethical Consideration

Ethical approval was obtained from the University of Cape Town Faculty of Health Sciences Human Research Ethics Committee (Protocol Number: 220/2024) and the Arba Minch University Institutional Research Ethics Review Board Office (Protocol Number: IRB/2320/2024).

## Informed Consent

Waiver for informed consent was granted.

## Figure and Tables

- Figure 1: CONSORT diagram: screening, exclusion, and final study population
- Table 1: Baseline characteristics of surgical patients by intervention and control group
- Table 2: Compliance rate with the recommended ERAS triple interventions
- Table 3: Postoperative complications by Clavien-Dindo classification across study groups
- Table 4: Factors influencing hospital length of stay, description of intervention and control groups

## Supplementary files

- Appendix 1: ERAS Trial collaborators

## References

1. Meara JG, Leather AJM, Hagander L, Alkire BC, Alonso N, Ameh EA, et al. Global Surgery 2030: Evidence and solutions for achieving health, welfare, and economic development. Vol. 386, The Lancet. Lancet Publishing Group; 2015. p. 569–624.

2. Weiser TG, Haynes AB, Molina G, Lipsitz SR, Esquivel MM, Uribe-Leitz T, et al. Estimate of the global volume of surgery in 2012: an assessment supporting improved health outcomes. The Lancet [Internet]. 2015 Apr 27 [cited 2025 Jun 16];385:S11. Available from: https://www.thelancet.com/action/showFullText?pii=S0140673615608066

3. International Surgical Outcomes Study group T. Global patient outcomes after elective surgery: prospective cohort study in 27 low-, middle-and high-income countries The International Surgical Outcomes Study group †. Br J Anaesth [Internet]. 2016 [cited 2025 Feb 8];117:601–9. Available from: www.isos.org.uk/docu

4. Biccard BM, Madiba TE, Kluyts HL, Munlemvo DM, Madzimbamuto FD, Basenero A, et al. Perioperative patient outcomes in the African Surgical Outcomes Study: a 7-day prospective observational cohort study. Lancet [Internet]. 2018 Apr 21 [cited 2023 Dec 7];391(10130):1589–98. Available from: https://pubmed.ncbi.nlm.nih.gov/29306587/

5. Kelly CM, Starr N, Raykar NP, Yorlets RR, Liu C, Derbew M. Provision of surgical care in Ethiopia: Challenges and solutions. Glob Public Health [Internet]. 2018 Nov 2 [cited 2025 Feb 8];13(11):1691–701. Available from: https://www.tandfonline.com/doi/abs/10.1080/17441692.2018.1436720

6. Burssa D, Teshome A, Iverson K, Ahearn O, Ashengo T, Barash D, et al. Safe Surgery for All: Early Lessons from Implementing a National Government-Driven Surgical Plan in Ethiopia. World J Surg [Internet]. 2017 Dec 1 [cited 2023 Apr 19];41(12):3038–45. Available from: https://link.springer.com/article/10.1007/s00268-017-4271-5

7. Ljungqvist O, Scott M, Fearon KC. Enhanced Recovery After Surgery: A Review. JAMA Surg [Internet]. 2017 Mar 1 [cited 2024 Feb 5];152(3):292–8. Available from: https://jamanetwork.com/journals/jamasurgery/fullarticle/2595921

8. Enhanced Recovery after Surgery (ERAS®) for Gastrointestinal Surgery in Africa: A Systematic Review and Meta-Analysis – Clinical Surgery Journal (ISSN 2767-0023) [Internet]. [cited 2023 Sep 10]. Available from: https://clinicalsurgeryjournal.com/article/1000115/enhanced-recovery-after-surgery-eras-for-gastrointestinal-surgery-in-africa-a-systematic-review-and-meta-analysis

9. McQueen K, Oodit R, Derbew M, Banguti P, Ljungqvist O. Enhanced Recovery After Surgery for Low– and Middle-Income Countries. World J Surg [Internet]. 2018 Apr 1 [cited 2023 Sep 10];42(4):950–2. Available from: https://link.springer.com/article/10.1007/s00268-018-4481-5

10. Kifle F, Belay E, Kifleyohanes T, Demissie B, Galcha D, Mulye B, et al. Adherence to Enhanced Recovery After Surgery (ERAS) With Bellwether Surgical Procedures in Ethiopia: A Retrospective Study. World J Surg [Internet]. 2025 [cited 2025 Apr 1]; Available from: https://onlinelibrary.wiley.com/doi/full/10.1002/wjs.12526

11. Turner L, Shamseer L, Altman DG, Weeks L, Peters J, Kober T, et al. Consolidated standards of reporting trials (CONSORT) and the completeness of reporting of randomised controlled trials (RCTs) published in medical journals. Cochrane Database of Systematic Reviews [Internet]. 2012 Nov 14 [cited 2025 Feb 9];2013(1). Available from: https://www.cochranelibrary.com/cdsr/doi/10.1002/14651858.MR000030.pub2/full

12. Fitsum Kifle. Pan African Clinical Trials Registry. 2025 [cited 2025 Feb 10]. Assessing the effectiveness of implementing ERAS protocols on hospital length of stay and patient outcomes in Bellwether surgical procedures in Ethiopia: Randomized Pragmatic Cluster Trial. Available from: https://pactr.samrc.ac.za/TrialDisplay.aspx?TrialID=33295

13. Kifle F, Iverson KR, Belay E, Buno Teko E, Dawit A, Deneke A, et al. Towards Establishing a National Perioperative Quality Improvement Network in LMICs: Implementation Experiences From Ethiopia. Annals of Surgery Open [Internet]. 2024 Aug 19 [cited 2024 Sep 4];5(3):e480. Available from: https://journals.lww.com/aosopen/fulltext/2024/09000/towards_establishing_a_national_perioperative.32.aspx

14. Oodit R, Biccard BM, Panieri E, Alvarez AO, Sioson MRS, Maswime S, et al. Guidelines for Perioperative Care in Elective Abdominal and Pelvic Surgery at Primary and Secondary Hospitals in Low–Middle-Income Countries (LMIC’s): Enhanced Recovery After Surgery (ERAS) Society Recommendation. World Journal of Surgery 2022 46:8 [Internet]. 2022 May 31 [cited 2023 Apr 19];46(8):1826–43. Available from: https://link.springer.com/article/10.1007/s00268-022-06587-w

15. Course: About SAFE | SAFE Online [Internet]. [cited 2025 Feb 11]. Available from: https://safe-anaesthesia.org/course/view.php?id=19

16. Donner A, Klar N (2000) – The James Lind Library The James Lind Library [Internet]. [cited 2025 Feb 13]. Available from: https://www.jameslindlibrary.org/donner-a-klar-n-2000/

17. Killip S, Mahfoud Z, Pearce K. What Is an Intracluster Correlation Coefficient? Crucial Concepts for Primary Care Researchers. The Annals of Family Medicine [Internet]. 2004 May 1 [cited 2025 Feb 14];2(3):204–8. Available from: https://www.annfammed.org/content/2/3/204

18. Debas SA, Chekol WB, Zeleke ME, Mersha AT. Delayed ambulation in adult patients after major abdominal surgery in Northwest Ethiopia: a multicenter prospective follow up study. Sci Rep [Internet]. 2025 Dec 1 [cited 2025 May 20];15(1):1–11. Available from: https://www.nature.com/articles/s41598-025-97933-0

19. Zhang L, Wu Q, Wang X, Zhu X, Shi Y, Wu CJ. Factors impacting early mobilization according to the Enhanced Recovery After Surgery guideline following gastrointestinal surgery: A prospective study. Geriatr Gerontol Int [Internet]. 2024 Feb 1 [cited 2025 May 20];24(2):234–9. Available from: /doi/pdf/10.1111/ggi.14799

20. Hu Y, McArthur A, Yu Z. Early postoperative mobilization in patients undergoing abdominal surgery: A best practice implementation project. JBI Database System Rev Implement Rep [Internet]. 2019 Dec 1 [cited 2025 May 20];17(12):2591–611. Available from: https://journals.lww.com/jbisrir/fulltext/2019/12000/early_postoperative_mobilization_in_patients.17.aspx

21. Baluku M, Bajunirwe F, Ngonzi J, Kiwanuka J, Ttendo S. A Randomized Controlled Trial of Enhanced Recovery after Surgery Versus Standard of Care Recovery for Emergency Cesarean Deliveries at Mbarara Hospital, Uganda. Anesth Analg [Internet]. 2020 Mar 1 [cited 2023 Nov 30];130(3):769–76. Available from: https://journals.lww.com/anesthesia-analgesia/fulltext/2020/03000/a_randomized_controlled_trial_of_enhanced_recovery.29.aspx

22. Kifle F, Kenna P, Daniel S, Maswime S, Biccard B. A scoping review of Enhanced Recovery After Surgery (ERAS), protocol implementation, and its impact on surgical outcomes and healthcare systems in Africa. Perioperative Medicine 2024 13:1 [Internet]. 2024 Aug 2 [cited 2025 Feb 28];13(1):1–12. Available from: https://perioperativemedicinejournal.biomedcentral.com/articles/10.1186/s13741-024-00435-2

23. Riad AM, Barry A, Knight SR, Arbaugh CJ, Haque PD, Weiser TG, et al. Perioperative optimisation in low– and middle-income countries (LMICs): A systematic review and meta-analysis of enhanced recovery after surgery (ERAS). J Glob Health [Internet]. 2023 [cited 2025 Mar 2];13:04114. Available from: https://pmc.ncbi.nlm.nih.gov/articles/PMC10546475/

24. Biccard BM, du Toit L, Lesosky M, Stephens T, Myer L, Prempeh AB, et al. Enhanced postoperative surveillance versus standard of care to reduce mortality among adult surgical patients in Africa (ASOS-2): a cluster-randomised controlled trial. Lancet Glob Health [Internet]. 2021 Oct 1 [cited 2025 May 20];9(10):e1391–401. Available from: https://www.thelancet.com/action/showFullText?pii=S2214109X21002916

25. Stokes SM, Scaife CL, Brooke BS, Glasgow RE, Mulvihill SJ, Finlayson SRG, et al. Hospital Costs Following Surgical Complications: A Value-driven Outcomes Analysis of Cost Savings Due to Complication Prevention. Ann Surg [Internet]. 2022 Feb 1 [cited 2025 Apr 30];275(2):E375–81. Available from: https://journals.lww.com/annalsofsurgery/fulltext/2022/02000/hospital_costs_following_surgical_complicationsa.42.aspx

26. Shrime MG, Bickler SW, Alkire BC, Mock C. Global burden of surgical disease: An estimation from the provider perspective. Lancet Glob Health [Internet]. 2015 Apr 27 [cited 2025 Feb 19];3(S2):S8–9. Available from: http://www.thelancet.com/article/S2214109X14703845/fulltext

27. Kamarajah S, Ademuyiwa AO, Atun R, Cieza A, Agyei F, Ghosh D, et al. Health systems strengthening through surgical and perioperative care pathways: a changing paradigm. BMJ Glob Health [Internet]. 2024 Nov 7 [cited 2025 May 20];9(Suppl 4):15058. Available from: https://gh.bmj.com/content/9/Suppl_4/e015058

28. Currie A, Burch J, Jenkins JT, Faiz O, Kennedy RH, Ljungqvist O, et al. The impact of enhanced recovery protocol compliance on elective colorectal cancer resection: Results from an international registry. Ann Surg [Internet]. 2015 Jun 1 [cited 2025 May 8];261(6):1153–9. Available from: https://journals.lww.com/annalsofsurgery/fulltext/2015/06000/the_impact_of_enhanced_recovery_protocol.20.aspx

29. Interactive Audit – ERAS® Society [Internet]. [cited 2025 May 8]. Available from: https://erassociety.org/interactive-audit/

