## Appendix 1_ERAS trial collaborators for "Targeted ERAS implementation for postoperative care after Bellwether procedures in Africa: A pragmatic cluster-randomized trial from Ethiopia"

Title: Improving postoperative recovery through targeted ERAS implementation in Africa: A pragmatic cluster-randomized trial from Ethiopia

### Authors

ERAS trial writing committee

^1^Fitsum Kifle Belachew (PhD), ^2^Peniel Kenna Dula (MSc), ^2^Ermiyas Belay Woldesenbet, ^2,3^Betelehem Mulye (MD), ^4^Desta Galcha (MD), ^2^Kalkidan Kifle (BSc), ^5^Dagmawi Dagne (MD), ^6^Megbar Desalegn Mekonnen (MD), ^2,7^Kokeb Desta Belihu (MSc), ^2,7^ Tewodros Kifleyohannes (MD), ^8^ Brook Demisse (MD), ^9^Abiy Dawit (MPH), ^10^ Salome Maswime (PhD), ^10,11^ Bruce Biccard (PhD) ^10,11^ **on behalf of NaPQIN ERAS trial collaborators/investigators**

**Affiliations**

^1^Global Surgery Division, Department of Surgery, Faculty of Health Sciences, University of Cape Town, Cape Town, South Africa

^2^ Global Partner for Improving Surgical System, Network for Perioperative and Critical Care (GPISS-N4PCc), Addis Ababa, Ethiopia.

^3^ Quality directorate, St. Peter comprehensive specialized Hospital, Addis Ababa, Ethiopia.

^4^ Assistant Professor, Department of surgery, Arba Minch University, South-Ethiopia.

^5^Department of General Surgery, Alert comprehensive specialized Hospital, Addis Ababa, Ethiopia

^6^Department of Surgery, Debre Markos Hospital Comprehensive Specialized, Debre Markos, Ethiopia.

^7^School of Medicine, Asrat Wolde Yes Health Science Campas, Debrebrehane University, Debrebrehane, Ethiopia.

^8^Medical Service Hospital and Diagnostic Desk, Federal Ministry of Health, Addis Ababa, Ethiopia

^9^Alert comprehensive specialized Hospital, Addis Ababa, Ethiopia.

^10^Global Surgery Division, Department of Surgery, Faculty of Health Sciences, University of Cape Town, Cape Town, South Africa.

^11^ Department of Anaesthesia and Perioperative Medicine, Groote Schuur Hospital, University of Cape Town, Cape Town, Western Cape, South Africa.

**NaPQIN ERAS trial collaborators/investigators**

^1^ Yordanos Damtew Tefera, ^2^ Beimnet Hailu Gebreegziabher, ^3^ Terefu Tsegaye Shitaye, ^3^ Tekalign Taye Tawede, ^4^ Abere Sinshaw, ^5^ Melkamu Siferih, ^6^ Adugnaw Bogale Worku, ^6^Zemenu Abiye Kassie, ^6^Mohammed Seid Ali, ^7^ Atalel Fentahun Awedew, ^7^ Moges Asnakew Wendm, ^8^Masresha Gebru Teklehaimanot, ^9^Aregawi Welay Abera, ^10^Shamill Eanga Helill, ^10^Helen Ashebir Haile, ^10^Netsanet Habte Gebre

^1^Data quality Analysis officer, Alert comprehensive specialized Hospital, Addis Ababa, Ethiopia.

^2^ Department of Surgery, Yekatit 12 Hospital Medical College, Addis Ababa, Ethiopia.

^3^Arbaminch General Hospital, Arba Minch, Ethiopia.

^4^ Department of Anesthesia, Debre Berhan Comprehensive Specialized Hospital, Debre Berhan, Ethiopia.

^5^ Debre Markos Comprehensive specialized hospital, Debre Markos, Ethiopia.

^6^ Mizan Tepi University Teaching Hospital, Mizan Tepi, Ethiopia

^7^ Debre Tabor University, Debre Tabor, Ethiopia

^8^ Impact Africa, Mekelle, Ethiopia.

^9^ Mekelle General Hospital, Mekelle, Ethiopia.

^10^ Department of Anesthesia, College of health Science, Wolkite university Comprehensive specialized Hospital, Wolkite, Ethiopia.
